# Estimation of the actual incidence of coronavirus disease (COVID-19) in emergent hotspots: The example of Hokkaido, Japan during February–March 2020

**DOI:** 10.1101/2020.04.24.20077800

**Authors:** Andrei R. Akhmetzhanov, Kenji Mizumoto, Sung-mok Jung, Natalie M. Linton, Ryosuke Omori, Hiroshi Nishiura

## Abstract

Following the first report of coronavirus disease 2019 (COVID-19) in Sapporo City, Hokkaido Prefecture, Japan on 14 February 2020, a surge of cases was observed in Hokkaido during February and March. As of 6 March, 90 cases were diagnosed in Hokkaido. Unfortunately, many infected persons may not have been recognized as cases due to having mild or no symptoms. We therefore estimated the actual number of COVID-19 cases in (i) Hokkaido Prefecture and (ii) Sapporo City using data on cases diagnosed outside these areas. The estimated cumulative incidence in Hokkaido as of 27 February was 2297 cases (95% confidence interval [CI]: 382, 7091) based on data on travelers outbound from Hokkaido. The cumulative incidence in Sapporo City as of 28 February was estimated at 2233 cases (95% CI: 0, 4893) based on the count of confirmed cases within Hokkaido. Both approaches resulted in similar estimates, indicating higher incidence of infections in Hokkaido than were detected by the surveillance system. This quantification of the gap between detected and estimated cases can help inform public health response as it provides insight into the possible scope of undetected transmission.

## 1 Introduction

In December 2019 a cluster of 41 patients with atypical pneumonia of unknown etiology was reported in Wuhan City, China [1,2]. The number of atypical pneumonia cases then rapidly increased in Wuhan in early January and cases began appearing across China and in other countries [3,4]. The cause of the pneumonia was recognized as a newly emerged coronavirus, called severe acute respiratory syndrome coronavirus 2 (SARS-CoV-2), and the disease it causes was named coronavirus disease 2019 (COVID-19). Cases of COVID-19 have now been reported in more than 120 countries worldwide [5], and the WHO declared it a pandemic on 11 March 2020. In the present paper, we examine the recent COVID-19 hotspot of Hokkaido Prefecture, Japan, and offer two approaches for estimation of actual incidence in the prefecture.

The first imported case of COVID-19 infection in Hokkaido was confirmed on 28 January 2020. The second case was reported two weeks later on 14 February 2020 in a male Sapporo (the capital of Hokkaido) resident in his 50s. At present, there is no evidence that the first imported case was linked to any subsequent cases, indicating either a possible hidden chain of transmissions that led to later incidence or that some imported cases were missed by the surveillance system. A surge of cases with history of travel to the Sapporo Snow Festival—held between 31 January and 11 February 2020—was detected two weeks after the festival ended, indicating transmission likely occurred during the event. From late February to early March the cumulative case count in Hokkaido tripled, from 30 cases reported by 24 February to 90 cases reported by 6 March. Among these cases, two were fatal. In addition, six cases were exported to other parts of Japan, and two cases were exported to Thailand and Malaysia. The increasing number of cases and exportation of infections signaled the urgent need to increase containment efforts within Hokkaido, and we were motivated to derive real-time estimates of the actual incidence in Hokkaido and Sapporo, respectively, by accounting for these underascertained cases in order to better understand the risk of further spread within and exportation from Hokkaido.

Control of COVID-19 spread is complicated by various factors, including: nonspecific clinical symptoms—especially during the early phase of infection [6–8], a relatively high proportion of pre-symptomatic and asymptomatic infections [9–11], multiple possible routes of transmission—including aerosol, direct contact, and fecal-oral transmission [8, 12, 13]—as well as the moderate to high potential of superspreading events generated by a small proportion of cases [14, 15]. Additionally, the incubation period can vary widely [16,17] and re-occurrence of the disease after primary infection is possible [18]. Nonetheless, the transmission dynamics of COVID-19 in the community can be roughly grasped by exploring epidemiological indicators such as the number of confirmed infections in a given location, or the number of exported cases.

Clinical manifestation of disease begins with nonspecific symptoms (similar to seasonal influenza) and symptoms remain mild for most persons (if any are developed). The disease may resolve into life-threatening onset of acute respiratory distress in a small fraction of cases [19]. Containment efforts for COVID-19 are underway, but it appears that the number of confirmed cases worldwide represents only a fraction of all infections, and the actual number of cases including mild and asymptomatic infections may be as large as four-fold [11, 20]. Hence, the derivation of reliable estimates of the actual number of cases is urgently needed to conduct risk assessments of the spread of COVID-19 and inform policies related to containment and mitigation [21]. This is especially important for newly emerging hotspots of the disease such as Iran, Italy, South Korea, and Japan [4,22].

Here, we estimate the incidence in: (i) Hokkaido Prefecture, using the number of detected cases among international and domestic travelers outbound from Hokkaido, and (ii) Sapporo City, using the number of confirmed cases within different subprefectures of Hokkaido. We build our framework on two key assumptions. First, we regard the outbreak to be in its initial stage, when the first cases in Sapporo emerged and had just expanded to other areas. In such an instance, we can still disentangle locally acquired infections from the imported cases in each subprefecture. Second, we assume that the travel volume between subprefectures in Hokkaido can be well approximated by models of commuters [23–25]. Assuming that our time period of February to March 2020 covers the initial phase of the epidemic in these areas, we presume that both estimates should be close to each other, because Sapporo was the epicenter for COVID-19 spread within Hokkaido.

## 2 Methods

### 2.1 Epidemiological data

Three sources of data were leveraged to estimate the actual incidence of COVID-19 in Hokkaido. First, cases exported domestically and internationally from Hokkaido were identified from government reports. The domestically exported cases were Kumamoto (1 case), Nagano (1 case) and Chiba (1 case) prefecture residents [26–28]. The internationally exported cases were reported from Malaysia (1 case) and Thailand (2 cases) [29, 30]. Considering that the two Thailand cases are husband and wife and one case is asymptomatic, one infection may have been the result of household transmission as similarly described in [31] rather than exported from Japan. Second, the total volume of air passengers from all airports in Hokkaido to Thailand and Malaysia over the two-month period of January and February 2019 were retrieved from the International Air Transport Association (IATA), and includes 9,349 passengers to Malaysia and 45,137 to Thailand. Lastly, the population size of Hokkaido was obtained from the Hokkaido Prefectural Government [32].

Calculation of COVID-19 incidence in Sapporo was based on cases of non-Sapporo residents who (i) became infected in Sapporo and were diagnosed outside of Sapporo or (ii) had no information on their source of infection (i.e. unknown link). Cases who were infected outside of Sapporo by Sapporo residents were excluded from the analysis, as they did not represent true exportation of infection from Sapporo by a local case. For example, in one instance a Sapporo resident travelled to Kitami City in Okhotsk subprefecture and caused secondary infections among Kitami residents. These Kitami residents were excluded from our analysis as they were infected outside of Sapporo. However, if a case from Kitami was to travel to Sapporo, have likely been infected there, and then be subsequently diagnosed in Kitami, they would eligible for our analysis. Data on cases reported through the end of February 2020, including dates of illness onset, subprefecture of diagnosis, travel history to Sapporo, and epidemiologic linkage to other confirmed cases were retrieved from the websites of government entities in Hokkaido.

The estimate of the fraction of commuters between subprefectures was obtained by exploring survey data of daytime and a nighttime population sizes in Sapporo [33]. By subtracting the daytime population size from the nighttime population size and dividing the difference by the nighttime population, we obtained a value of 3.7% [33], reflecting the change in population size of Sapporo due to commuting. However, this estimate accounts for movement of individuals both within Ishikari subprefecture and between subprefectures. In contrast, our model requires the average estimate between all subprefectures of Hokkaido. Therefore, we set this parameter at 1% and performed a sensitivity analysis wherein we varied the fraction of commuters between 0.5% and 4%.

### 2.2 Reporting delay between illness onset and confirmation

The time interval from illness onset *S_i_* to report of case confirmation *R_i_* was extracted for each confirmed case *i*. The likelihood function was doubly interval-censored [16, 34]:

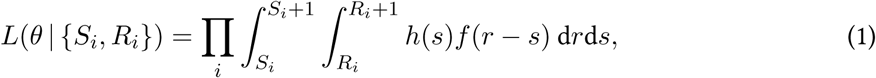

with *h*(*·*) as the probability distribution function (PDF) of illness onset time following a uniform distribution, and *f*(*·*) as the PDF of the reporting delay independent of *h*(*·*). We fit the distribution *f*(*·*) with parameters *θ* to gamma, lognormal, and Weibull distributions, and we selected the best-fit distribution by comparing widely acceptable information criterion (WAIC) values.

An alternative way is to implement the right truncated likelihood:

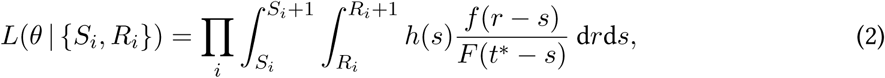

where *t^∗^* is the cut-off time at noon of 29 February 2020, *F* (*·*) is the cumulative distribution function of the distribution *f*(*·*). However, our analysis has shown that the mean delay was longer than two weeks and this was inconsistent with real observations. The reason is that the right truncation does not account for the effect of control measures implemented in the late February (see Discussion).

### 2.3 Estimating incidence in Hokkaido by using imported cases among travelers outbound from Hokkaido

We implemented a balance equation to predict incidence across all Hokkaido using the case count of air travelers from Hokkaido with international or domestic destinations [3,35–37]. Given the observed cumulative count of exported cases *C* we write our balance equation as follows:

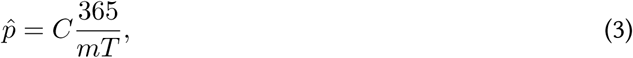

where *m* is total volume of passengers to the corresponding destination, *T* is the infectious period, approximated from the observed virus shedding period, 5.0 days [38]. The estimated incidence in Hokkaido is given by 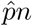, where *n* is the catchment population size for the international airport in Hokkaido. The cumulative incidence was estimated using maximum likelihood and the 95% confidence intervals were calculated using profile likelihood using data generated following the binomial sampling process among travelers from Hokkaido.

### 2.4 Estimating the incidence in Sapporo cases imported by other areas of Hokkaido

Let *C_j_* be the cumulative number of cases who had no link to Sapporo or any other local cluster (see case classification in the Method section) and were detected in subprefecture *j*. Following a linear regression model with negative binomial link [39] we obtain:

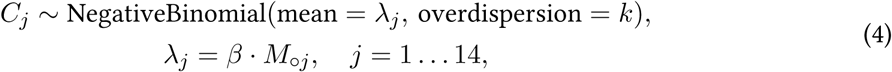

where *β* is the regression coefficient, *k* is the overdispersion parameter, and *M_◦j_* is the predicted commuter flow between Sapporo and the various subprefectures of Hokkaido. Because Sapporo is a part of Ishikari subprefecture, we define Ishikari subprefecture as the area that excludes Sapporo (Figure 1B). The flow *M_◦j_* was predicted using the uniform selection and radiation models of human movement [40, 41], as described below.

**Figure 1.**
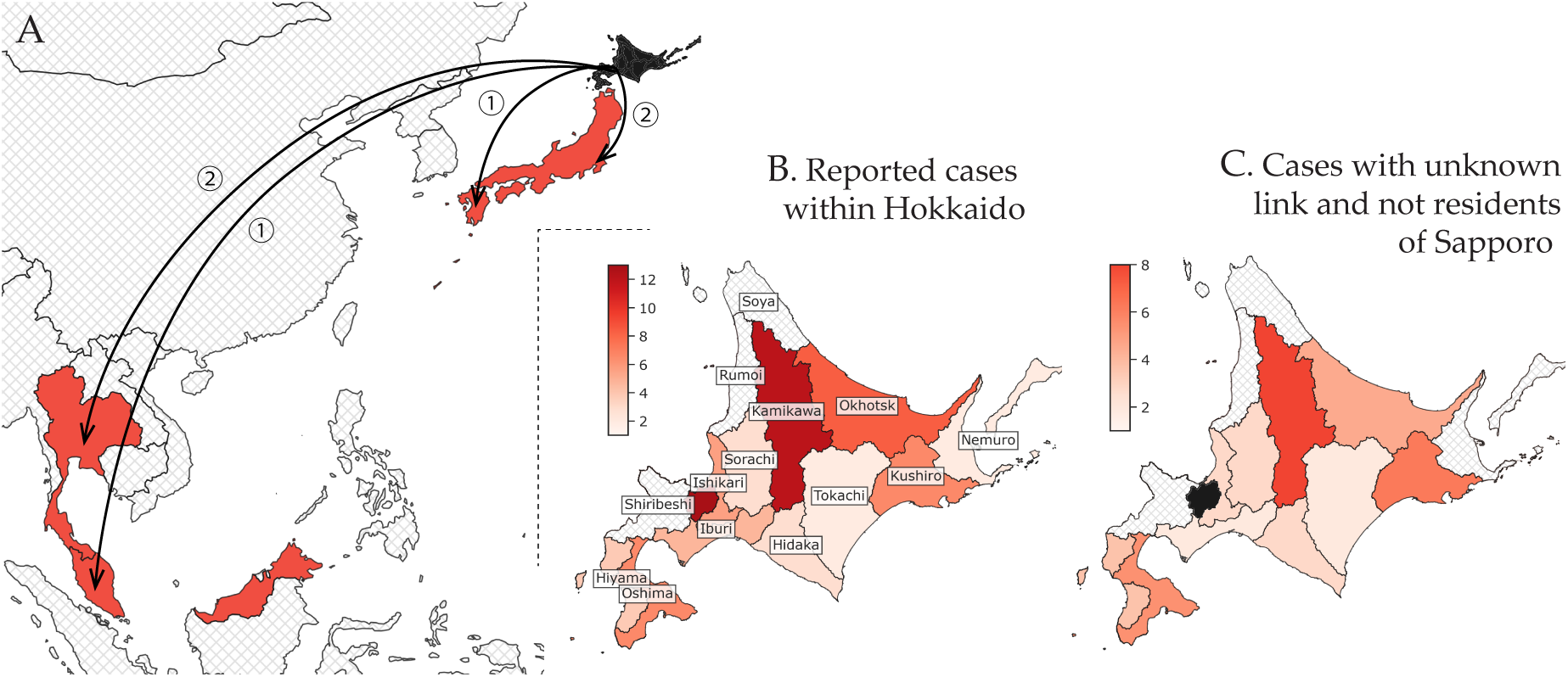
Geographical distribution of confirmed cases linked to Hokkaido by subprefecture of diagnosis as of 29 February 2020. (**A**) shows international and domestic cases (excluding Hokkaido). Hokkaido is shaded in black, whereas affected countries are depicted in red. The circled numbers indicate the total count of cases for each import destination. (**B**) shows all cases confirmed in Hokkaido. The labels indicate the names of the subprefectures of Hokkaido. (**C**) shows only cases included in our analysis: non-residents of Sapporo with unknown links either with or without travel history to Sapporo. Ishikari subprefecture was separated into two subregions: Sapporo City and outside Sapporo City. Hatched areas in grey indicate subprefectures with zero counts.

The final estimate for the cumulative incidence in Sapporo was obtained by extending (4) to the entire population of Sapporo as follows:

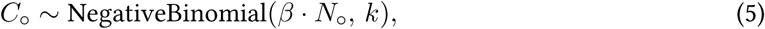

where *N_◦_* is the population size of Sapporo.

### 2.5 Modeling commuter movement between subprefectures of Hokkaido

Two single-parameter models of human movement were employed to predict the rate of commuting between Sapporo and the various subprefectures of Hokkaido—a radiation model and a model of uniform selection. Both emphasize the attractiveness of large population centers [23,24,42]. The radiation model incorporates the distance metric between the origin and destination while accounting for their population sizes. The uniform selection model [41] predicts the commuting volume using the formula:

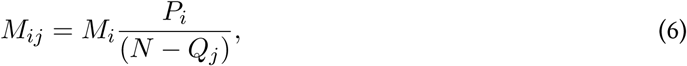

where *P_i_* is the population size at the origin *i*, *Q_j_* is the population size at the destination *j*, *N* is the total population of Hokkaido. The only parameter *M_i_* defines the scaling parameter of the ongoing number of commuters from the source subprefecture and equals to the proportion of commuters multiplied on the population size *P_i_* [42]. The radiation model [40] defines a commuting volume similarly to (6):

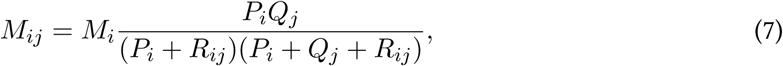

where *R_ij_* denotes the total population within a radius around two centers *P_i_* and *Q_j_*. Both matrices *M_ij_* (6)–(7) were forced to be symmetric in our simulations.

An aggregated dataset of population sizes and central point coordinates for each subprefecture was used to estimate the commuting rates between Sapporo and the subprefectures of Hokkaido while leveraging the R package “movement” [25]. The package required a fraction of the commuting individuals to be a single input parameter for the function. Due uncertainty of this value, we performed a sensitivity analysis with reference to the daytime and nighttime populations of Sapporo [33].

The derived estimates were then used to validate our assumption of linearity between the number of reported COVID-19 infections in each subprefecture and the daily commuting rate [39]. The number of infections *C_i_* in subprefecture *i* is described by a negative binomial distribution with mean *λ_i_* linearly proportional to the travel volume *x_i_*: *λ_i_* = *β·x_i_*, and a dispersion parameter *k*. In this case, the variance of the distribution takes the form: 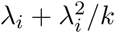. The regression parameter *β* is regarded as independent of the region. Using the fit of the predicted travel volume and observed case counts, we selected the best-fit model between (6)–(7) based on the difference in WAIC values and with lower value regarded as better.

The data were processed using R version 3.6.2 and Python version 3.6.10. The Markov chain Monte Carlo (MCMC) simulations were performed in Stan (cmdStan version 2.22.1 [43]) for estimation of the delay distribution, and in PyMC3 version 3.8 [44] for all other estimates. All code is freely available at the github repository: http://github.com/aakhmetz/Covid19IncidenceHokkaidoFeb2020.

## 3 Results

### 3.1 Epidemiological situation

The first case was reported in Hokkaido on 28 January 2020. A month later on 28 February 2020, the total count of confirmed cases reached 65, with 13 cases reported by Sapporo, and another 51 case reported by 11 of the 14 subprefectures of Hokkaido. The place of diagnosis of the first case was unspecified. Although the initial cluster of cases was linked to the Sapporo Snow Festival, the overall geographic distribution of COVID-19 cases appeared to be widespread. By 28 February Japan had also been notified of three cases diagnosed in Thailand and Malaysia who were believed to have been infected in Hokkaido, as well as three cases domestic cases with exposures in Hokkaido that were reported by other prefectures (Table 1, Figure 1A).

**Table 1.** Exportation events and estimated incidence in Hokkaido by date of report. CI, confidence interval (the 95% CI was derived from profile likelihood); NA, not available. The estimated incidence is updated by the function of the date of report. The estimated incidence for the latest available reporting date (27 February 2020) depends on whether both cases 4 and 5 acquired their infection in Hokkaido (#) or whether one was infected by the other (##).

| Case number | Diagnosis location | Date of illness onset | Date of report | Cumulative count | Estimated incidence in Hokkaido (95% CI) |
| --- | --- | --- | --- | --- | --- |
| 1 | Kumamoto | 15-Feb-20 | 22-Feb-20 | 2 | 24 (4, 74) |
| 2 | Chiba | 16-Feb-20 |  |  |  |
| 3 | Nagano | 20-Feb-20 | 25-Feb-20 | 3 | 36 (9, 93) |
| 4-5 | Thailand | 20-Feb-20<br>NA | 26-Feb-20 | 6 | 3446 (857, 8931) <sup>#</sup><br>2298 (382, 7091) <sup>##</sup> |
| 6 | Malaysia | 25-Feb-20 | 27-Feb-20 |  |  |

A surge of cases was observed at the end of February (Figure 2A). In the first half of February, peaks of reported cases by date of illness onset were separated by 3–4 days on average, consistent with estimates of the serial interval [45]. The epidemic curve peaked around 18 February 2020, but the decline in cases which was seen later can be explained by the delay in reporting. The delay distribution fitted with the gamma distribution had the mean of 7.9 days (95% CI: 6.9, 9.0) and standard deviation of 4.2 days (95% CI: 3.3, 5.2). The 95th percentile was 15.6 days (95% CI: 13.4, 18.8) implying that cases with illness onset in the last two weeks of February were likely to be underascertained.

**Figure 2.**
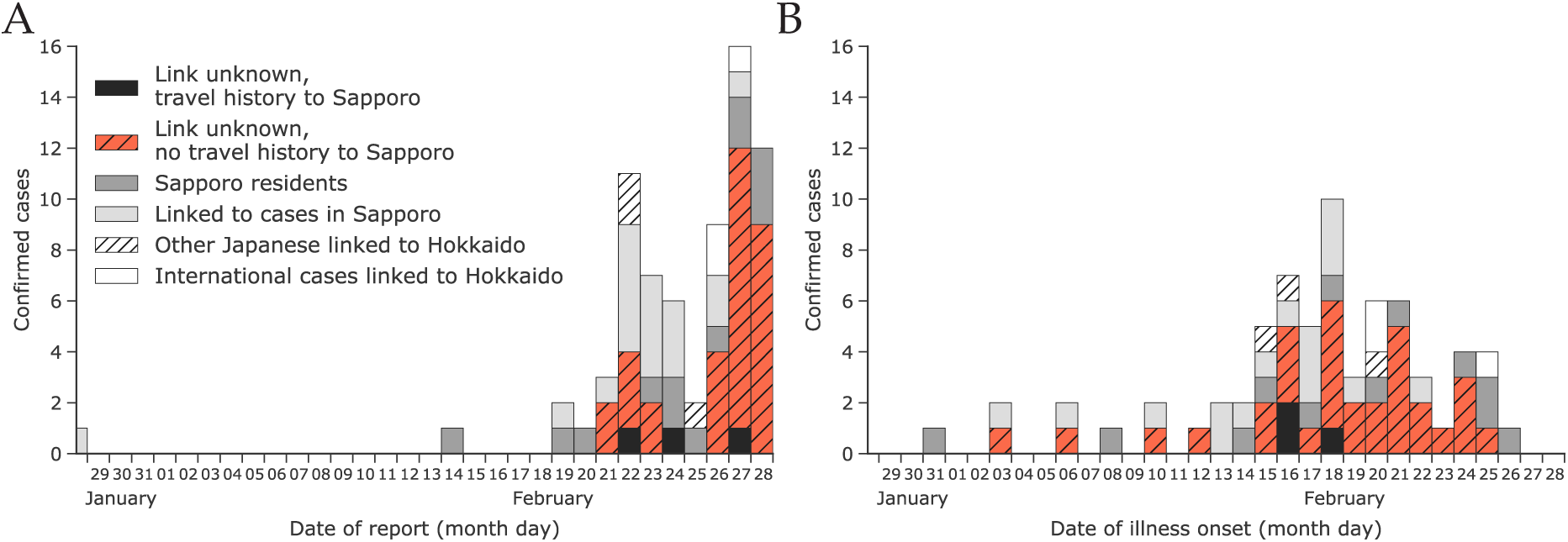
Epidemic curves by date of report (A) and date of illness onset (B) as of 29 February 2020 for confirmed cases among Japanese nationals that were linked to Hokkaido. The total number of cases are characterized according to the legend shown in (A).

### 3.2 Estimated incidence in Hokkaido using confirmed cases diagnosed outside Hokkaido

The first three cases diagnosed in early February outside of Hokkaido indicated a low incidence of COVID-19 in Hokkaido with an estimated upper bound (95th percentile) of fewer than 100 cases. Our estimate of the cumulative incidence in Hokkaido as of 25 February 2020 is dependent on whether the transmission within the infected couple who travelled to Thailand was acquired in the community or within the household, and we estimated incidence in Hokkaido to be higher in the former scenario, at 3446 cases (95% CI: 857, 8931), compared to the latter, with 2297 cases (95% CI: 382, 7091), see Figure 3A.

**Figure 3.**
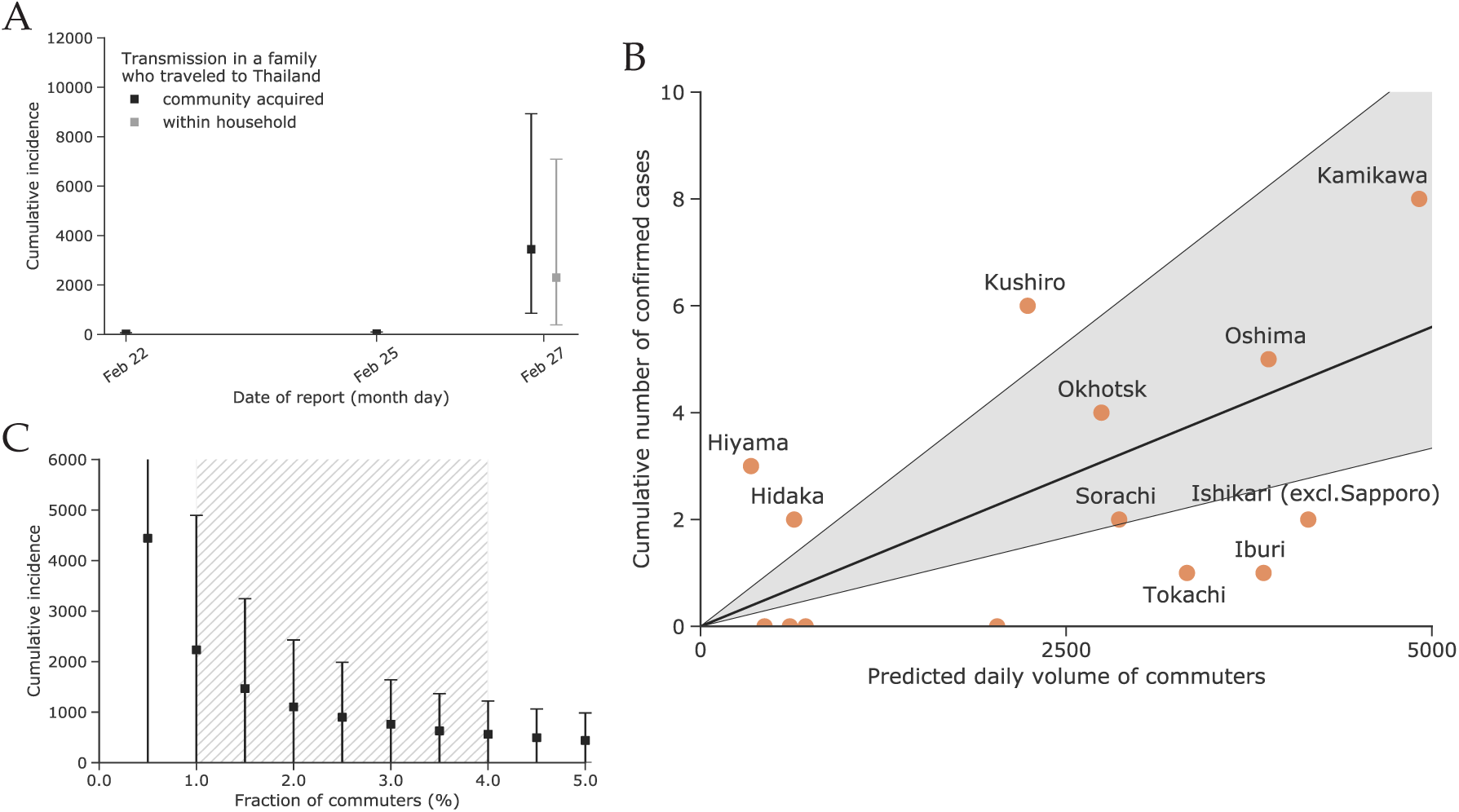
(**A**) Estimated cumulative incidence of coronavirus disease 2019 (COVID-2019) in Hokkaido using the count of imported cases either internationally or domestically outbound from Hokkaido. (**B**) Modeled daily volume of commuters and observed incidence of COVID-19 by subprefecture of diagnosis. The four subprefectures with zero case counts are Rumoi, Soya, Nemuro, and Shirebeshi (from left to right). (**C**) Estimated cumulative incidence in Sapporo as of 29 February with a varying fraction of commuters. The plausible range between 0.5% and 4% is indicated by hatched grey area. The square points in (A) and (C) indicate the estimated mean, whereas the error bars indicate the 95% confidence intervals.

### 3.3 Estimated incidence in Sapporo using confirmed cases diagnosed within Hokkaido

A total of 34 Hokkaido cases were included in our model (Figure 1B). We fit the radiation and uniform selection models to the population data for each subprefecture based on their centroids (Supplementary figure 1). The resulting fit to the observed incidence of COVID-19 cases showed that the uniform selection model performed better than the radiation model (WAIC values were 60.2 vs 68.3, respectively). Figure 3B shows the fit using the model with uniform selection. If we assume the fraction of commuters to be at 1%, the mean estimated incidence in Sapporo was 2233 cases (95% CI: 0, 4893) as of 28 February 2020. Variation in the fraction of commuters between 0.5% and 4% results in estimated incidence between 4440 (95% CI: 0, 9687) and 563 (95% CI: 0, 1221) for the 0.5% and 4%, respectively (Figure 3C).

## 4 Discussion

In our analysis of the reporting delay, we did not implement right truncation of the likelihood because the low case count (by date of illness onset) seen in late February through the beginning of March was likely due to the effect of interventions in Hokkaido rather than delays in reporting. In support of this assumption, there were fewer cases reported between 28 February and 6 March compared to the week of 21–27 February (cf. Figure 2 and Supplementary figure 2). Right truncation should therefore be used with caution when it is unclear whether recent incidence is increasing or decreasing.

Although we tracked the spread of the disease in Hokkaido and we were able to distinguish between locally acquired and travel associated infections, the surveillance systems in the subprefectures of Hokkaido may not be sensitive enough to detect cases with milder symptoms. This could explain why we obtained a lower estimate of the incidence in Sapporo using cases diagnosed within Hokkaido compared to analyzing the outbound flow of travelers from Hokkaido. Use of data on travel volume of commuters and tourists within Hokkaido could be beneficial for fitting the travel volume to the observed incidence rather than adopting a model of human movement (Figure 3B). However, publicly available data on train, bus, and private car travel in Hokkaido were inconsistent and scarce, and we were unable to use them for this study. Nevertheless, our results demonstrate that the use of mathematical models of human movement at the scale of estimating the travel volume between administrative subunits of a given region is a promising alternative to use of observed transportation data. Previously, this has been also demonstrated by other researchers in assessing the risk of spread of yellow fever in Angola [23] or during epidemics in resource poor settings [24].

In addition, surveillance among travelers at international borders may detect infections not found through standard surveillance systems due to implementation of additional detection methods [46,47]. As shown by other researchers [39,48] the linear regression fit of the travel volume among international travelers to observed disease incidence represents an elegant way to distinguish areas that are able to well detect new infections from those that are likely to miss some fraction of cases. We applied the same approach to compare the performance of the various subprefectures of Hokkaido, as shown in Figure 3B. Subprefectures that fall within the 95% CI of the linear regression line are considered to have surveillance systems that succeed in detecting COVID-19 infections within their jurisdiction. Subprefectures that fall below the 95% CI of the linear regression line may not have surveillance systems that are sensitive enough to detect all imported cases [39]. The four subprefectures with no reported cases are Rumoi, Soya, Nemuro, and Shirebeshi. The first three are distant and less connected to Sapporo compared to, for example, Kamikawa or Oshima, which contain the second and third largest cities of Hokkaido, Asahikawa and Hakodate (Figure 1B). However, Shirebeshi is located next to Ishikari/Sapporo and includes the relatively large city of Otaru, though the lack of reports in Shiribeshi may be explained by our use of place of diagnosis over place of residence for determining the subprefecture assigned to cases. This was done because the specific place of residence was not always reported. We suspect that some cases from Shirebeshi subprefecture were diagnosed in Ishikari subprefecture.

Our estimate of the incidence in Sapporo is in the range of 1,000 to 10,000 cases, which is similar to early estimates of the incidence in Wuhan City, China that used data on the first cases among international travelers [3, 49]. Assuming our estimates are correct, a substantial proportion of these infected persons likely had only a mild or asymptomatic course of disease [9, 17] and therefore did not seek medical care and were not detected by the surveillance system.

Preliminary analysis of the offspring distribution among infected individuals in the detected clusters around Japan has shown that approximately 80% of cases do not produce secondary infections (i.e., their reproduction number is equal to zero) [50]. Because most of the clusters have been linked to the known cases and could be traced back in time, we argue that there is still a window of opportunity for containment of the disease and only some small fraction of the infections may drive the epidemic [14]. We agree with other studies, e.g. [51,52], that a successful strategy for control of COVID-19 lays in strict movement restriction, avoiding social gatherings, and intense investigation effort in contact tracing.

## Data Availability

Data on COVID-19 confirmed cases, including dates of illness onset, subprefecture of diagnosis, travel history to Sapporo, and epidemiologic linkage to other confirmed cases were retrieved from the websites of government entities in Hokkaido.

http://github.com/aakhmetz/Covid19IncidenceHokkaidoFeb2020

## Author Contributions

A.R.A., K.M. and H.N. conceived the study and participated in the study design. S.-m.J., K.M. and N.M.L. assisted in data collection. A.R.A. and K.M. analyzed the data. A.R.A., N.M.L., and K.M. drafted the manuscript. All authors edited the manuscript and approved the final version.

---

**Supplementary figure 1.**
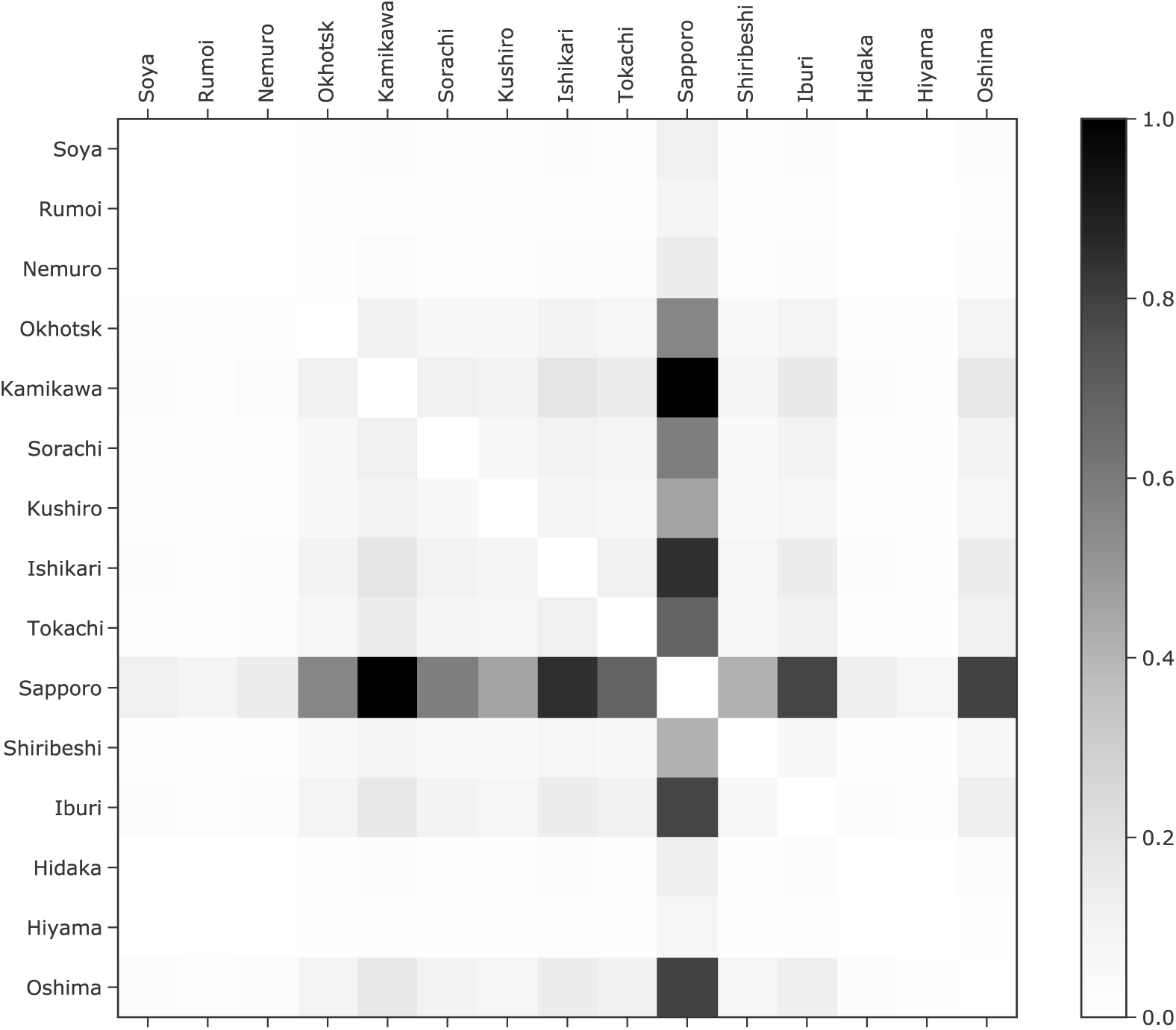
Reconstructed connectivity matrix between subprefectures of Hokkaido. Ishikari subprefecture was separated into two subregions: within Sapporo and outside of Sapporo. The grey-shaded heatmap indicates the relative intensity of travel volume. The resulting connectivity matrix was forced to be symmetric.

**Supplementary figure 2.**
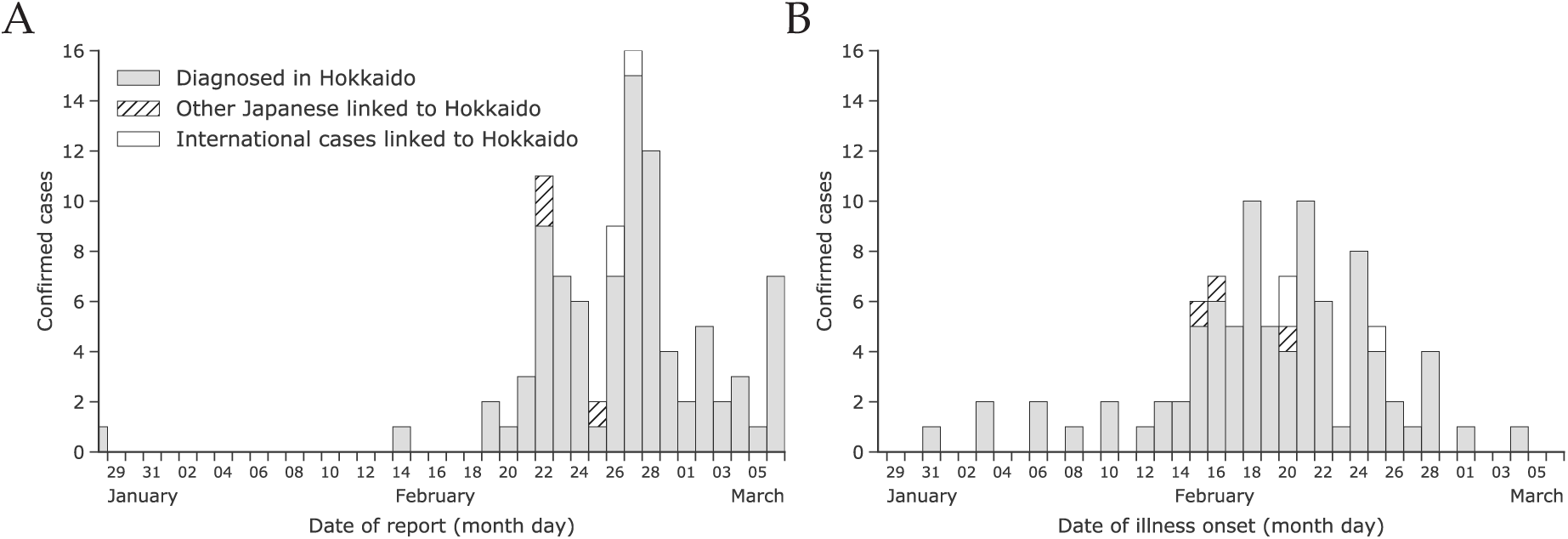
Epidemic curves by date of confirmation (A) and date of illness onset (B) as of 6 March 2020 for confirmed cases among Japanese nationals linked to Hokkaido.

